# Sustained impact of 10-valent pneumococcal conjugate vaccine on invasive pneumococcal disease in Kenya, 2011-2022

**DOI:** 10.1101/2024.04.22.24306163

**Authors:** E Wangeci Kagucia, Brian M Nyamwaya, Gerald Ongayo, Mary Kaniu, Samuel Sang, Ruth Lucinde, Angela Karani, Donald Akech, Fredrick Odiwuor, Christine Mataza, Collins Tabu, Neema Mturi, Siti Ndaa, Caroline Mulunda, Timothy Etyang, Nadia Aliyan, Amek Nyaguara, Shirine Voller, Christian Bottomley, Laura Hammitt, Ifedayo Adetifa, J Anthony G Scott

**Author notes:** Contact information Corresponding author: E. Wangeci Kagucia, PhD KEMRI-Wellcome Trust Research Programme Department of Epidemiology and Demography, P. O. Box 230-80108, Kilifi, Kenya.

## Abstract

**Background:** There are only a few long-term PCV impact assessments in sub-Saharan Africa, and these have been confined to settings using a 13-valent PCV. A 10-valent PCV was introduced in Kenya in 2011 with catchup vaccination among children aged <5 years in Kilifi. We evaluated the impact of PCV10 introduction in Kilifi through 2022.

**Methods:** Surveillance for IPD among residents of the Kilifi Health and Demographic Surveillance System was conducted at the Kilifi County Referral Hospital. Identification of pneumococcus isolated from blood or cerebrospinal fluid and pneumococcal serotyping were conducted according to WHO recommendations. Age– and serotype-specific incidence rate ratios, adjusted for pre-defined confounders (aIRRs), were used to compare annual IPD incidence in the pre-vaccine period to that in 2017-2019 (late post-vaccine) and 2020-2022 (COVID-19).

**Findings:** Compared to the pre-vaccine period, the incidence of vaccine serotype (VT) IPD among children aged <5 years was significantly lower in 2017-2019 (aIRR 0.14; 95%CI 0.04-0.49) and in 2020-2022 (aIRR 0.03; 95%CI 0.00-0.25). It also declined among older children and adults. The incidence of non-VT (NVT) IPD among children aged <15 years was higher during the post-vaccine period. All serotype IPD incidence declined across all age groups. Among individuals with NVT-IPD, serotypes included in new-generation PCVs accounted for about one-third and about one-half of disease among individuals aged <5 years and ≥5 years, respectively.

**Interpretation:** Despite potential waning of the effects of catchup vaccination during introduction, reductions in VT-IPD incidence were sustained through 12 years of PCV10 use. All serotype IPD incidence declined across all ages despite serotype replacement among children. New-generation PCVs may enhance IPD control.

## Introduction

Studies in various settings globally have shown substantial impact of pneumococcal conjugate vaccines (PCVs) in reducing invasive pneumococcal disease (IPD) among children targeted for vaccination (i.e., direct effects) as well as among unvaccinated individuals through the indirect effects of the vaccines.^1–5^ Before introduction of PCVs, children in sub-Saharan Africa bore a disproportionate burden of pneumococcal disease.^6^ Despite this, the published evidence on the impact of PCV on IPD in sub-Saharan Africa^7–20^ is disproportionately small. In addition, most of the available evidence from sub-Saharan Africa describes the short-term impact of PCV, i.e., within ≤5 years of PCV introduction. The available yet limited long-term evidence on PCV impact in sub-Saharan Africa^10,12,17^ describes the impact of 13-valent PCV (PCV13). In The Gambia and Malawi, vaccine effectiveness against vaccine serotype IPD among children aged <5 years ranged from 74-92% within six to eight years of PCV13 use. Vaccine effectiveness ranged from 42-80% against IPD caused by all serotypes in the same age group. However, there is no long-term evidence of the impact of 10-valent PCVs (PCV10) within sub-Saharan Africa.

PCV10 (Synflorix^TM^) – targeting serotypes 1, 4, 5, 6B, 7F, 9V, 14, 18C, 19F, and 23F – was introduced into the national infant immunization program in Kenya in 2011. To accelerate the population level impact, PCV10 was introduced in Kilifi County with catchup vaccination among children aged <5 years, in contrast to the rest of the country where the catchup was restricted to children aged <12 months. By 2016 PCV10 use in Kilifi was associated with a 92% reduction in the incidence of IPD due to vaccine serotypes (VT-IPD) among children aged <5 years, 74% among those aged 5-14 years, and 81% among those ≥15 years of age. It was also associated with a 68% reduction in the incidence of all serotype IPD among children <5 years of age and a 53% reduction among children aged 5-14 years.^8^

Several important questions related to the long-term impact of PCV in Kenya remain. First, despite the remarkable reductions in the incidence of VT-IPD within six years of PCV use, the residual nasopharyngeal carriage prevalence of VT pneumococci among children in Kilifi was higher than expected. For example, among children aged <5 years, it was approximately 9% in Kilifi compared to <1% in the UK within six years of PCV introduction in each respective setting.^8,21^ Given that pneumococcal carriage is a precursor to invasive disease, these relatively high levels of residual VT carriage in Kilifi raise the theoretical concern of a mitigation in the impact of PCV10 on VT-IPD over time as the population level protection conferred by the catchup vaccination campaign wanes.

Another theoretical concern with long-term use of PCVs has been serotype replacement disease. In Kilifi, reductions in carriage of VT were completely negated by increases in carriage of non-VT (NVT), resulting in equilibration of all serotype pneumococcal carriage to pre-vaccine levels by 2016, five years after PCV10 introduction.^8^ In some settings, increases in non-vaccine serotype (NVT) disease have undermined reductions in VT-IPD among older age groups over the longterm.^22–26^

Third, the COVID-19 pandemic^27^ interrupted routine immunization systems globally in early 2020.^28^ Although immunization program operations began to recover pre-pandemic levels later in 2020, there remained concerns about the implications of missed vaccinations in the earlier part of the year. At the same time, the impact of COVID-19 control measures – such as movement restrictions, school closures and masking mandates – as well as that of the subsequent easing of these measures has not been well elucidated in Kenya and similar settings. In the UK for example, significant declines in IPD incidence were observed during the COVID-19 pandemic^29^ followed by substantial increases among children after COVID-19 restrictions were lifted.^30^

Finally, as Kenya begins to transition out of funding support from Gavi, the Vaccine Alliance,^31^ an understanding of the long-term impact of PCV will be important for justifying continued investments into the PCV program, as well as for optimizing PCV product selection in the context of new-generation PCVs which are less costly (e.g., 10-valent Pneumosil™) or which offer wider serotype coverage (e.g., 13-, 15– and 20-valent PCVs).

We sought to evaluate the long-term impact of 10-valent Synflorix™ on IPD, including during the COVID-19 period, in Kenya.

## Methods

### Study design and participants

The participants and design for this study have been described in detail elsewhere.^8,32^ Briefly, this was a before-after study involving individuals resident in Kilifi Health and Demography Surveillance System (HDSS)^33^ who were admitted to the Kilifi County Referral Hospital (KCRH). The IPD surveillance included all paediatric patients except those with trauma or admitted for elective surgery and all patients admitted to the adult ward with suspected invasive bacterial disease. In addition, vaccination coverage monitoring included a sample of 500 children in each of the following age groups, who were randomly selected from the Kilifi HDSS population register: 6 weeks – 11 months, 12-23 months, and 24-59 months.

Written informed consent was obtained from adults and guardians of children participating in the IPD surveillance and vaccination coverage monitoring. Ethical approval to conduct the study was granted by Kenya Medical Research Institute Scientific Ethics Review Unit (SSC 1433) and the Oxford Tropical Research Ethics Committee (30-10).

### Procedures

#### IPD surveillance

A blood culture sample was collected for eligible children and adults admitted at KCRH. A cerebrospinal fluid (CSF) sample was collected among children and adults with suspected meningitis. Blood was collected in BD BACTEC Peds Plus^TM^ and BD BACTEC^TM^ Plus Aerobic/F culture vials and processed using the automated BD BACTEC^TM^ FX blood culture system. CSF samples were cultured on blood and chocolate agar. As previously described,^8^ pneumococcal isolates were identified by optochin susceptibility and serotyped using latex agglutination and Quellung reaction. Confirmatory PCR testing was performed on a subset. Pneumococcal identification and serotyping were conducted in accordance with World Health Organization (WHO) recommendations.^34^ Serotypes included in Synflorix™ were classified as VT. Clinical care was provided by hospital staff in accordance with Kenya Ministry of Health guidelines.

#### Vaccination coverage monitoring

Through March 2018, vaccination coverage within the KHDSS was monitored using an electronic system that has been described previously.^35,36^ To obtain vaccination coverage estimates since 2019, we conducted a vaccination coverage survey in 2021.^37^ Vaccination status was ascertained through either review of the home-based vaccination record, or verbal report if the home-based vaccination record was not available. Health/ sociodemographic data was collected from parents/ guardians of the selected children. Study staff referred guardians of under– or un-vaccinated children to the nearest health facility for vaccination.

### Statistical analysis

#### IPD analysis

The pre-vaccine introduction period was defined as starting on 1 January 1999 for the IPD analysis among children <15 years and 1 January 2007 for the IPD analysis among individuals ≥15 years. It ended on 31 December 2022 for both analyses. The post-vaccine period was defined as 1 January 2012 to 31 December 2022. 2011 was excluded from the analysis since it was the year of PCV10 introduction.

To calculate age-stratified IPD incidence rates, the numerators were the annual number of IPD cases and the denominators were the mid-year population estimates from the KHDSS. Health worker strike periods, where surveillance was temporarily suspended, were excluded from the analysis. As data were overdispersed, negative binomial regression with the mid-year population as an offset term was used to estimate incidence rate ratios (IRRs) and associated 95% confidence intervals (95%CI) for each post-vaccine period compared to the pre-vaccine period. The post-vaccine periods were: early post-vaccine (2012-2016), late post-vaccine (2017-2019), and COVID-19 (2020-2022). Regression models were built for each age-group (i.e., <2 months, <5 years, 5-14 years, and ≥15 years) and serotype category (all serotype, vaccine serotype and non-vaccine serotype IPD). IRRs were adjusted for previously identified confounders,^8^ namely, year for the <2 months and <5 years age groups, and blood culture collection for the 5-14 years and ≥15 years age groups. To assess whether the impact of PCV10 was different across the various post-vaccine periods, a Wald test of the equality of the post-vaccine period coefficients was conducted for each age– and serotype-category regression.

#### Vaccination coverage analysis

Coverage with three doses of PCV10 was estimated for 2018 to 2021 using the sample included in the 2021 vaccination coverage survey. For each year, the denominator was the number of children who would have been aged 12-23 months by 15 July of the respective year (conditioned on the date of the vaccination coverage survey visit for 2021) and the numerator was the number of those children who had received three doses of PCV10 according to the home-based vaccination record. Exact 95%CIs were calculated for each annual vaccination coverage estimate.

Analyses were conducted using Stata version 15 (StataCorp LLC, College Station, TX).

## Results

The analysis included a total of 688 culture-confirmed IPD cases out of 69,575 admissions to KCRH. Most IPD cases occurred in the pre-vaccine period (558 of 688; 81.1%) and among children aged <5 years (460 of 688; 66.9%). The mean number of all-cause annual admissions among children aged <5 years was significantly higher in the pre-vaccine period than in the post-vaccine periods. In all age groups, the proportion of patients recommended for blood culture and the proportion with a blood culture done were lower in the post-vaccine periods than in the pre-vaccine period (Table 1).

**Table 1.** Descriptive characteristics of invasive pneumococcal disease (IPD) clinical surveillance among residents of the Kilifi Health and Demographic Surveillance System admitted to the Kilifi County Referral Hospital.

| Characteristic | Pre-vaccine period (1999-2010)*<br>n (%) | Early post-vaccine period (2012-2016)<br>n (%) | Late post-vaccine period (2017-2019)<br>n (%) | COVID-19 period (2020-2022)<br>n (%) | p-value |
| --- | --- | --- | --- | --- | --- |
| **Age <5 years** |  |  |  |  |  |
| Total admissions | 31422 | 6323 | 3066 | 3242 |  |
| Mean annual admissions | 2619 | 1265 | 1022 | 1081 | **<0.001** |
| Recommended for blood culture | 30098 (96%) | 5691 (90%) | 2764 (90%) | 2902 (90%) | **<0.001** |
| Blood culture collected† | 29234 (97%) | 5353 (94%) | 2491 (90%) | 2656 (92%) | **<0.001** |
| Culture-confirmed IPD | 401 | 34 | 14 | 11 |  |
| Age <24 months | 273 (68%) | 20 (59%) | 9 (64%) | 10 (91%) | 0.252 |
| Male | 229 (57%) | 23(68%) | 12 (86%) | 3 (27%) | **0.017** |
| Cultured from blood specimen | 390 (97%) | 34 (100%) | 13 (93%) | 9 (82%) | **0.039** |
| Cultured from CSF | 71 (18%) | 3 (9%) | 1 (7%) | 3 (27%) | 0.337 |
| Pneumococcal meningitis‡ | 78(19%) | 3 (9%) | 1 (7%) | 3 (27%) | 0.248 |
| HIV-infected§ | 25 (20%) | 4 (15%) | 2 (14%) | 0 (0%) | 0.163 |
| Died during the episode | 70 (17%) | 11 (32%) | 5 (36%) | 3 (27%) | 0.059 |
| **Age 5–14 years** |  |  |  |  |  |
| Total admissions | 5296 | 2060 | 951 | 887 |  |
| Mean annual admissions | 441 | 412 | 317 | 296 | 0.060 |
| Recommended for blood culture | 4483 (85%) | 1592 (77%) | 676 (71%) | 672 (76%) | **<0.001** |
| Blood culture collected† | 4200 (94%) | 1438 (90%) | 570 (84%) | 563 (84%) | **<0.001** |
| Culture-confirmed IPD | 127 | 22 | 9 | 4 |  |
| Male | 66(52%) | 10 (45%) | 6 (67%) | 0 (0%) | 0.156 |
| Cultured from blood specimen | 121 (95%) | 21 (95%) | 7 (78%) | 1 (25%) | **0.001** |
| Cultured from CSF | 31 (24%) | 2 (9%) | 2 (22%) | 2 (50%) | 0.195 |
| Pneumococcal meningitis‡ | 37 (29%) | 4 (18%) | 2 (22%) | 2 (50%) | 0.527 |
| HIV-infected§ | 7(23%) | 5(36%) | 2 (22%) | 0 (0%) | **0.008** |
| Died during the episode | 30 (24%) | 2 (9%)` | 1 (11%) | 0 (0%) | 0.339 |
| **Age ≥15 years** |  |  |  |  |  |
| Total admissions | 5307 | 5163 | 2826 | 3032 |  |
| Mean annual admissions | 1327 | 1033 | 942 | 1011 | 0.279 |
| Recommended for blood culture | 5025 (95%) | 4597 (89%) | 2415 (85%) | 2728 (90%) | **<0.001** |
| Blood culture collected† | 2084 (41%) | 1875 (41%) | 810 (34%) | 754 (28%) | **<0.001** |
| Culture-confirmed IPD | 30 | 26 | 7 | 3 |  |
| Male | 12(40%) | 11 (42%) | 2 (29%) | 3 (100) | 0.223 |
| Cultured from blood specimen | 29 (97%) | 23 (88%) | 6 (86%) | 2 (67%) | 0.155 |
| HIV-infected§ | 4 (33%) | 14 (64%) | 0 (0%) | 0 (0%) | **<0.001** |
| Died during the episode | 12 (40%) | 12 (46%) | 6 (86%) | 1 (33%) | 0.168 |

\*2007-2010 among individuals aged ≥15 years

Coverage with three doses of PCV10 among children aged 12-23 months increased from 85.5% (95%CI 84.7-86.3) in 2012 to 94.6% (95%CI 94.0-95.1) in 2016 then dipped to 88.6% (95%CI 87.8-89.4) in 2017.^36^ Coverage in 2018-2021, as estimated from the vaccination coverage survey, ranged from 76% to 92% (Table S1).

Among children aged <5 years, the annual incidence of VT-IPD declined significantly during late post-vaccine and COVID-19 periods (Figure 1). It was lower than in the pre-vaccine period by 86% (adjusted IRR [aIRR] 0.14; 95%CI 0.04-0.49) in 2017-2019 and by 97% (aIRR 0.03; 95%CI 0-0.25) in 2020-2022 (Tables 2 and S2). The annual incidence of VT-IPD also declined among children aged 5-14 years (aIRR 0.32; 95%CI 0.09-1.08) and among individuals aged ≥15 years in 2017-2019, while there were no cases of VT-IPD in these age groups in 2020-2022. Only one case of VT-IPD was detected among children aged <2 months in 2017-2019, equating to a slight increase in annual VT-IPD incidence (aIRR 1.28; 95%CI 0.09-19.01) compared to the pre-vaccine period (Tables 2 and S2).

**Figure 1.**
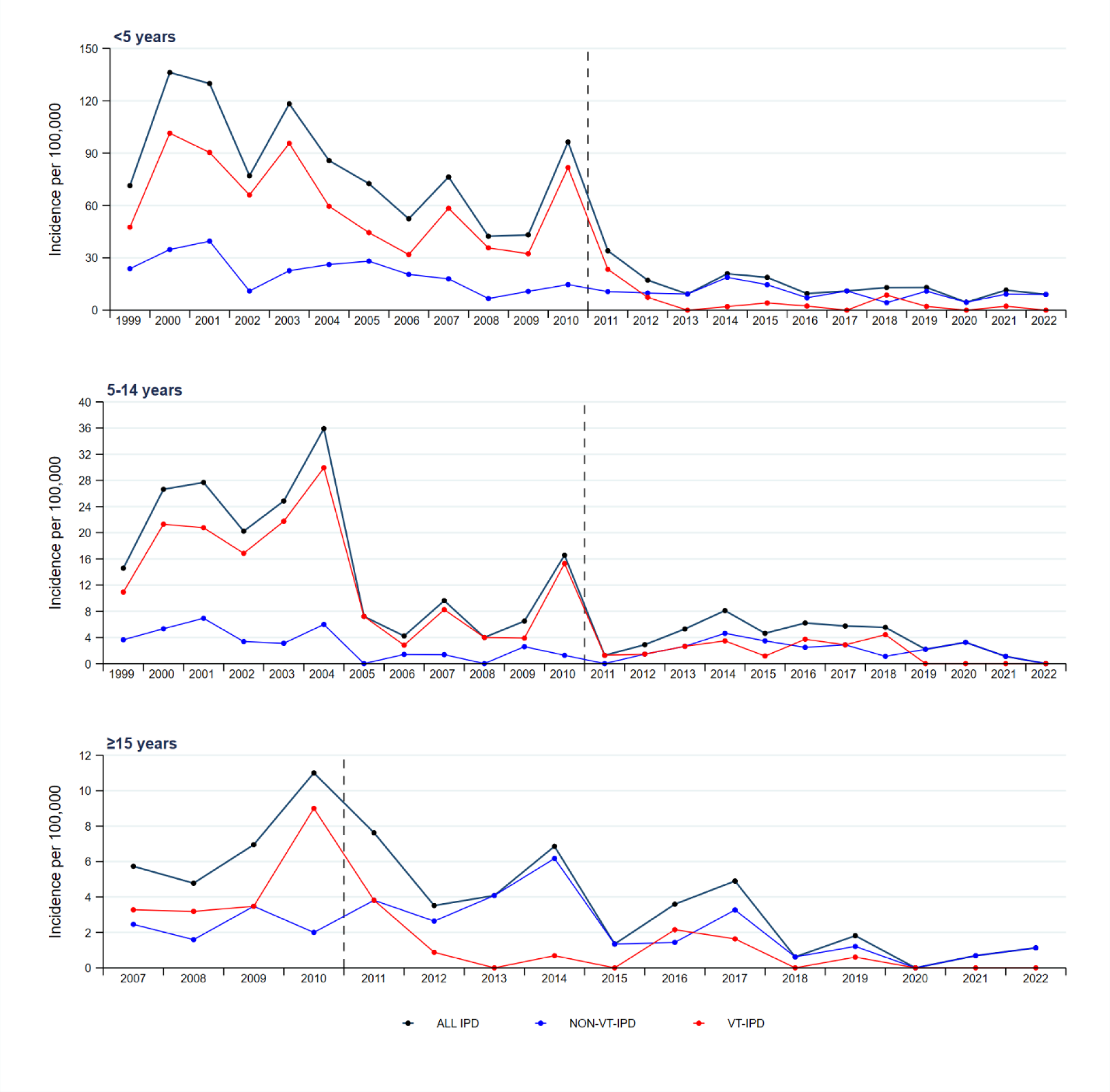
Incidence of vaccine serotype (VT), non-VT and all serotype invasive pneumococcal disease (IPD) among residents of the Kilifi Health and Demographic Surveillance system by age group, 1999-2022.

**Table 2.**
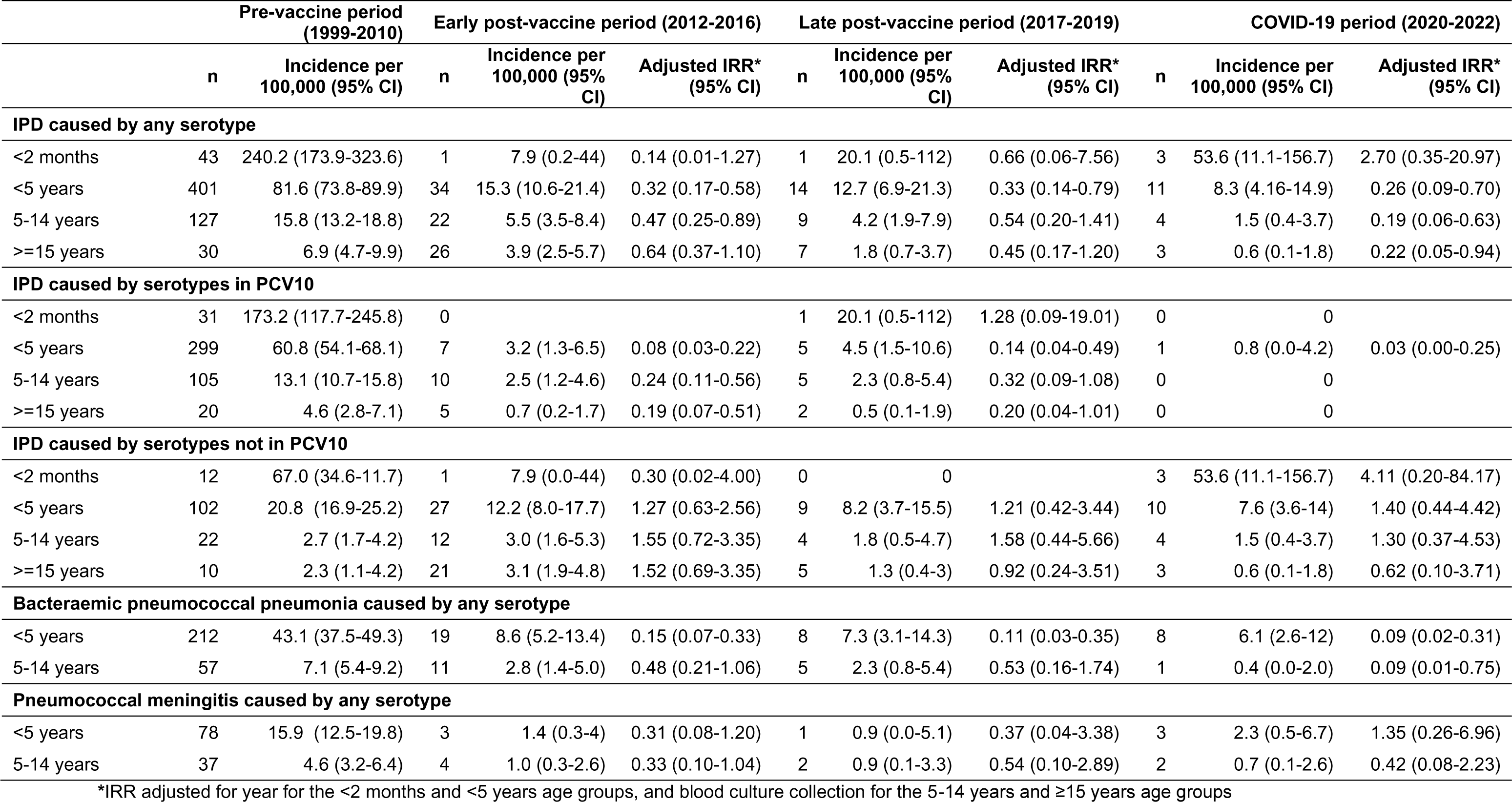
Incidence rates of invasive pneumococcal disease (IPD) and adjusted incidence rate ratios (IRR) comparing post-vaccine incidence rates to pre-vaccine incidence rates among residents of the Kilifi Health and Demographic Surveillance System, 1999-2022.

The annual incidence of NVT-IPD among children aged <5 years and 5-14 years was marginally higher in 2017-2019 and in 2020-2022 compared to the pre-vaccine period, with a net decrease in the incidence of all serotype IPD in these age groups in 2017-2019 (aIRR <5 years 0.33; 95%CI 0.14-0.79, aIRR 5-14 years 0.54; 95%CI 0.20-1.41) and in 2020-2022 (aIRR <5 years 0.26; 95%CI 0.09-0.70, aIRR 5-14 years 0.19; 95%CI 0.06-0.63; Tables 2 and S2). Among individuals aged ≥15 years, the annual incidence of NVT-IPD in 2017-2019 and 2020-2022 was lower than in the pre-vaccine period with overall reductions in the annual incidence of all serotype IPD. Among children aged <2 months there were no cases of NVT-IPD and a net decrease in the incidence of all serotype IPD in 2017-2019 compared to the pre-vaccine period. In 2020-2022 the annual incidence of NVT-IPD in this age group was higher than in the pre-vaccine period as was the annual incidence of all serotype IPD (Tables 2 and S2). The impact of PCV10 in 2017-2022 was no different from that in 2012-2016 (p-values for difference in coefficients >0.05) across all age groups, except for NVT-IPD and all serotype IPD among children aged <2 months in 2020-2022 (Table S2).

The annual incidence of bacteraemic pneumococcal pneumonia and among children aged <15 years in 2017-2019 and 2020-2022 continued to be lower than in the pre-vaccine period, not significantly different from the impact in the early post-vaccine period (Tables 2 and S2). Similarly, the impact of PCV10 on pneumococcal meningitis within the same age group in 2017-2022 was comparable to that in 2012-2016 (Tables 2 and S2).

Non-Synflorix™ serotypes 6A and 19A (included in 10-valent Pneumosil™ and 13-valent PCV [PCV13]) and serotype 3 (included in PCV13) accounted for approximately one-third or more of NVT-IPD cases among children aged <5 years. Non-Synflorix™ serotypes included in 20-valent PCV (PCV20) accounted for a little over half of NVT-IPD cases among individuals aged ≥5 years (Figure 2). No IPD cases due to 22F or 33F were detected during the post-vaccine period.

**Figure 2.**
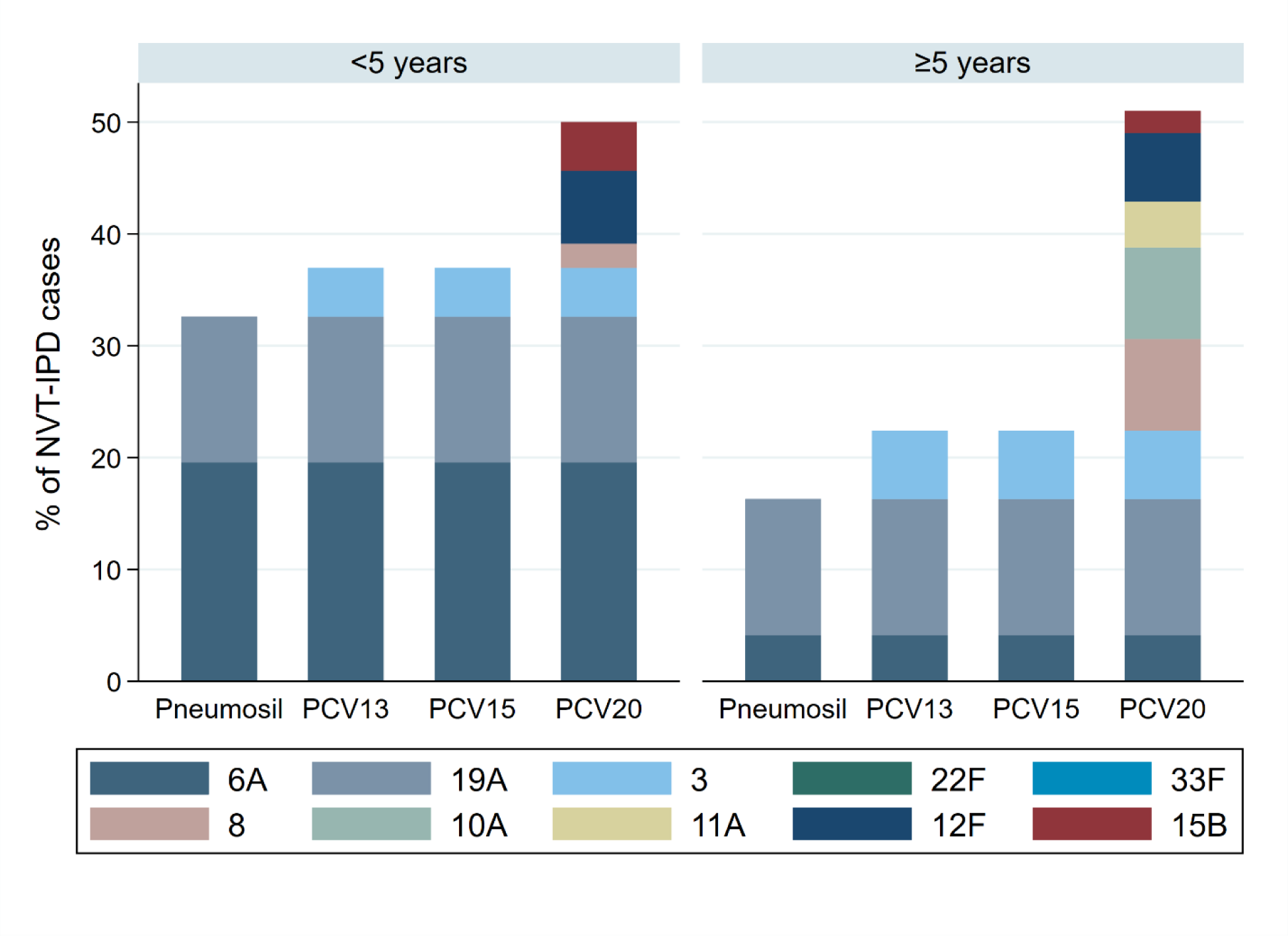
Distribution of non-vaccine serotype invasive pneumococcal disease (NVT-IPD) cases due to additional serotypes included in new-generation pneumococcal conjugate vaccines (PCVs) as a proportion of all NVT-IPD cases during the post-vaccine period, by age group.

## Discussion

We leveraged a 24-year-old population-based IPD surveillance programme within a lower-middle income setting to assess the long-term impact of Synflorix™. By 2017-2019, i.e., six to eight years after PCV10 introduction in Kenya, the annual incidence of VT-IPD among children aged <5 years and those aged 5-14 years was, respectively, 86% and 68% lower than the incidence prior to PCV10 introduction. During the same period, VT-IPD incidence was also lower than before vaccine introduction among older children and adults aged ≥15 years. By 2020-2022, including the period during the COVID-19 pandemic, the incidence of VT-IPD among children aged <5 years was 97% lower than during the pre-vaccine period and there were no cases of VT-IPD among children and adults aged ≥5 years. The impact in 2017-2019 and in 2020-2022 was not significantly different from that by 2016 when PCV10 use was associated with reductions in the incidence of VT-IPD of 92%, 76% and 81% among children aged <5 years, those aged 5-14 years and individuals aged ≥15 years, respectively.^8^ Protection against VT-IPD among children too young to be vaccinated also appeared to be sustained, with only one case of VT-IPD detected between 2017 and 2022. These findings indicate that any waning of the population-level impact of catchup vaccination among children aged <5 years alongside PCV10 introduction in Kilifi was not associated with a mitigation in the long-term impact of PCV10, aligning well with a previous modelling study that predicted a similar outcome.^38^

We also found increases in the incidence of NVT-IPD among children aged <15 years in 2017-2019 and in 2020-2022. However, these increases did not negate the PCV10-associated reductions in VT-IPD, resulting in sustained reductions of 46-81% in the incidence of all serotype IPD across all age groups. In settings similar to Kenya and with comparable duration of PCV use, there have been mixed findings on changes in the incidence of NVT-IPD after PCV introduction. In Malawi, the incidence of NVT-IPD was higher in the post-vaccine period than in the pre-vaccine period among children aged 1-4 years but not among older children and adults. In The Gambia, the incidence of NVT-IPD increased in the post-vaccine period compared to the pre-vaccine period among children aged 5-14 years but not among those aged <5 years.

Nevertheless, similar to Kenya, all serotype IPD declined among children in Malawi and The Gambia despite increases in NVT-IPD in some childhood age groups.^12,39^ Notably, the incidence of NVT-IPD among older children and adults aged ≥15 years in Kilifi was lower after long-term PCV10 use than in the pre-vaccine period. In contrast, in some settings, such as the UK, increases in non-vaccine serotype IPD incidence among older individuals, who may also be at higher risk for severe disease, have been equivalent to decreases in the incidence of VT-IPD, returning overall incidence of IPD to pre-vaccine introduction levels in the long-term.^22^ Consistent with findings from other long-term evaluations in sub-Saharan Africa,^12,39^ and other settings globally such as the US, we found no evidence of complete serotype replacement disease among older individuals.^10,12,40^

We found that serotypes 6A and 19A, included in the 10-valent Pneumosil™ and in PCV13, but not in Synflorix™, accounted for about one-third of NVT-IPD among children, indicating a role for new-generation PCVs in enhancing IPD control in Kenya. While no cases due to 22F and 33F were detected, culture-based identification of pneumococcus is acknowledged to be imperfect.^41^ Therefore, 15-valent PCV, which has recently been recommended for use in children and adults in the US and Europe,^42–44^ may also contribute to improved IPD control in Kenya. PCV20 has also been recently recommended for use in the US and Europe,^43,44^ albeit among adults. We found that eight of the 10 additional non-Synflorix™ serotypes included in PCV20 serotypes (i.e., except 22F and 33F) accounted for a little over half of NVT-IPD among older children and adults in Kilifi. Though the epidemiology of pneumococcal pneumonia in Kenya remains poorly understood, the NVT-IPD data may provide important insights into the contribution of these serotypes to adult pneumococcal disease and the potential role for PCV20 in adult disease control particularly among the vulnerable, e.g., the elderly and immunocompromised.

The impact of PCV10 on the annual incidence of VT-IPD among children aged <5 years during the early PCV10 use period (i.e., 2012-2016, a 92% reduction) was not appreciably different from that during the COVID-19 period (i.e., 2020-2022, a 97% reduction). We have previously shown that the COVID-19 pandemic did not interrupt infant immunization uptake in Kilifi,^37^ making this finding plausible. There were no cases of VT-IPD among children and adults aged ≥5 years in 2020-2022 and therefore any differences in PCV10 impact in the older age groups could not be estimated. However, there were no statistically significant differences in the magnitude of the impact of PCV10 on NVT-IPD in the early PCV10 use period *vs* during the COVID-19 period and no consistent trend in the change in effect sizes across the older age groups. Therefore, it is unlikely that the COVID-19 pandemic had a sustained impact on pneumococcal transmission within our setting.

Our analysis was limited by the small number of IPD cases in the post-vaccine period, increasing the uncertainty around the effect sizes in some strata. Nevertheless, the consistency in the direction of impact over the various post-vaccine periods lends credence to the findings. In addition, we observed differences in some surveillance characteristics over time. To minimize bias, we adjusted the analyses for previously identified confounders.

There are only a few long-term PCV impact assessments in sub-Saharan Africa, and these have been confined to settings using a 13-valent PCV. Nevertheless, long-term PCV impact assessments remain important for informing future PCV policy. Despite relatively high residual nasopharyngeal carriage vaccine serotypes, we found that the impact of Synflorix™ in Kenya was sustained over more than a decade. This finding supports continued investment in PCVs by the Government of Kenya and partners, particularly as Kenya approaches graduation from Gavi support. The low incidence of VT-IPD suggests minimal risk for a switch to a reduced dose schedule in Kenya, though this will be informed by emerging evidence on reduced dose PCV trials in settings such as The Gambia.^45^ The NVT-IPD serotype distribution suggests that new-generation PCVs have the potential to enhance IPD control among children and adults. In late 2022, Kenya transitioned to use of the 10-valent Pneumosil™, which differs from Synflorix™ by two serotypes; Pneumosil™ includes serotypes 6A and 19A in place of serotypes 4 and 18C. A theoretical risk for this product switch is a rebound in carriage of and IPD due to serotypes 4 and 18C. As more affordable and higher valency PCVs become available, there is a need for ongoing population-based surveillance for IPD to generate evidence on the impact of PCV program changes on IPD epidemiology, which can then be used to inform future policy decisions in Kenya and in other sub-Saharan African settings.

## Data Availability

All data produced in the present study are available upon reasonable request to the authors.

## References

1. Feikin DR, Kagucia EW, Loo JD, et al. Serotype-specific changes in invasive pneumococcal disease after pneumococcal conjugate vaccine introduction: a pooled analysis of multiple surveillance sites. PLoS Med 2013; 10(9): e1001517.

2. Tsaban G, Ben-Shimol S. Indirect (herd) protection, following pneumococcal conjugated vaccines introduction: A systematic review of the literature. Vaccine 2017; 35(22): 2882–91.

3. de Oliveira LH, Camacho LA, Coutinho ES, et al. Impact and Effectiveness of 10 and 13-Valent Pneumococcal Conjugate Vaccines on Hospitalization and Mortality in Children Aged Less than 5 Years in Latin American Countries: A Systematic Review. PLoS One 2016; 11(12): e0166736.

4. Cohen O, Knoll M, K. OB, et al. Pneumococcal conjugate vaccine (PCV) Review of Impact Evidence (PRIME): Summary of findings from systematic review. Geneva, Switzerland, 2017.

5. Hanquet G, Krizova P, Valentiner-Branth P, et al. Effect of childhood pneumococcal conjugate vaccination on invasive disease in older adults of 10 European countries: implications for adult vaccination. Thorax 2019; 74(5): 473–82.

6. Wahl B, O’Brien KL, Greenbaum A, et al. Burden of Streptococcus pneumoniae and Haemophilus influenzae type b disease in children in the era of conjugate vaccines: global, regional, and national estimates for 2000-15. Lancet Glob Health 2018; 6(7): e744–e57.

7. Bigogo GM, Audi A, Auko J, et al. Indirect Effects of 10-Valent Pneumococcal Conjugate Vaccine Against Adult Pneumococcal Pneumonia in Rural Western Kenya. Clin Infect Dis 2019; 69(12): 2177–84.

8. Hammitt LL, Etyang AO, Morpeth SC, et al. Effect of ten-valent pneumococcal conjugate vaccine on invasive pneumococcal disease and nasopharyngeal carriage in Kenya: a longitudinal surveillance study. Lancet 2019; 393(10186): 2146–54.

9. Mackenzie GA, Hill PC, Jeffries DJ, et al. Effect of the introduction of pneumococcal conjugate vaccination on invasive pneumococcal disease in The Gambia: a population-based surveillance study. Lancet Infect Dis 2016; 16(6): 703–11.

10. Mackenzie GA, Hill PC, Jeffries DJ, et al. Impact of the introduction of pneumococcal conjugate vaccination on invasive pneumococcal disease and pneumonia in The Gambia: 10 years of population-based surveillance. The Lancet Infectious Diseases 2021; 21(9): 1293–302.

11. Mackenzie GA, Hill PC, Sahito SM, et al. Impact of the introduction of pneumococcal conjugate vaccination on pneumonia in The Gambia: population-based surveillance and case-control studies. Lancet Infect Dis 2017; 17(9): 965–73.

12. Bar-Zeev N, Swarthout TD, Everett DB, et al. Impact and effectiveness of 13-valent pneumococcal conjugate vaccine on population incidence of vaccine and non-vaccine serotype invasive pneumococcal disease in Blantyre, Malawi, 2006&#x2013;18: prospective observational time-series and case-control studies. The Lancet Global Health 2021; 9(7): e989–e98.

13. von Gottberg A, de Gouveia L, Tempia S, et al. Effects of vaccination on invasive pneumococcal disease in South Africa. N Engl J Med 2014; 371(20): 1889–99.

14. Kaboré L, Ouattara S, Sawadogo F, et al. Impact of 13-valent pneumococcal conjugate vaccine on the incidence of hospitalizations for all-cause pneumonia among children aged less than 5 years in Burkina Faso: An interrupted time-series analysis. International Journal of Infectious Diseases 2020; 96: 31–8.

15. Soeters HM, Kambiré D, Sawadogo G, et al. Impact of 13-Valent Pneumococcal Conjugate Vaccine on Pneumococcal Meningitis, Burkina Faso, 2016–2017. The Journal of Infectious Diseases 2019; 220(Supplement_4): S253–S62.

16. Kambiré D, Soeters HM, Ouédraogo-Traoré R, et al. Early impact of 13-valent pneumococcal conjugate vaccine on pneumococcal meningitis-Burkina Faso, 2014-2015. The Journal of infection 2018; 76(3): 270–9.

17. Njuma Libwea J. A., Fletcher M, Koki Ndombo P, et al. Impact of 13-valent pneumococcal conjugate vaccine on laboratory-confirmed pneumococcal meningitis and purulent meningitis among children ˂5 years in Cameroon, 2011–2018. PLOS ONE 2021; 16(4): e0250010.

18. Faye PM, Sonko MA, Diop A, et al. Impact of 13-Valent Pneumococcal Conjugate Vaccine on Meningitis and Pneumonia Hospitalizations in Children aged <5 Years in Senegal, 2010–2016. Clinical Infectious Diseases 2019; 69(Supplement_2): S66–S71.

19. Nhantumbo AA, Weldegebriel G, Katsande R, et al. Surveillance of impact of PCV-10 vaccine on pneumococcal meningitis in Mozambique, 2013 – 2015. PLoS One 2017; 12(6): e0177746.

20. Gatera M, Uwimana J, Manzi E, et al. Use of administrative records to assess pneumococcal conjugate vaccine impact on pediatric meningitis and pneumonia hospitalizations in Rwanda. Vaccine 2016; 34(44): 5321–8.

21. van Hoek AJ, Sheppard CL, Andrews NJ, et al. Pneumococcal carriage in children and adults two years after introduction of the thirteen valent pneumococcal conjugate vaccine in England. Vaccine 2014; 32(34): 4349–55.

22. Ladhani SN, Collins S, Djennad A, et al. Rapid increase in non-vaccine serotypes causing invasive pneumococcal disease in England and Wales, 2000-17: a prospective national observational cohort study. Lancet Infect Dis 2018; 18(4): 441–51.

23. Regev-Yochay G, Katzir M, Strahilevitz J, et al. The herd effects of infant PCV7/PCV13 sequential implementation on adult invasive pneumococcal disease, six years post implementation; a nationwide study in Israel. Vaccine 2017; 35(18): 2449–56.

24. Galanis I, Lindstrand A, Darenberg J, et al. Effects of PCV7 and PCV13 on invasive pneumococcal disease and carriage in Stockholm, Sweden. European Respiratory Journal 2016; 47(4): 1208–18.

25. Mahmud SM, Sinnock H, Mostaço-Guidolin LC, Pabla G, Wierzbowski AK, Bozat-Emre S. Long-term trends in invasive pneumococcal disease in Manitoba, Canada. Hum Vaccin Immunother 2017; 13(8): 1884–91.

26. D’Ancona F, Caporali MG, Del Manso M, et al. Invasive pneumococcal disease in children and adults in seven Italian regions after the introduction of the conjugate vaccine, 2008-2014. Epidemiol Prev 2015; 39(4 Suppl 1): 134–8.

27. Cucinotta D, Vanelli M. WHO Declares COVID-19 a Pandemic. Acta Biomed 2020; 91(1): 157–60.

28. Shet A, Carr K, Danovaro-Holliday MC, et al. Impact of the SARS-CoV-2 pandemic on routine immunisation services: evidence of disruption and recovery from 170 countries and territories. Lancet Glob Health 2022; 10(2): e186–e94.

29. Amin-Chowdhury Z, Aiano F, Mensah A, et al. Impact of the Coronavirus Disease 2019 (COVID-19) Pandemic on Invasive Pneumococcal Disease and Risk of Pneumococcal Coinfection With Severe Acute Respiratory Syndrome Coronavirus 2 (SARS-CoV-2): Prospective National Cohort Study, England. Clin Infect Dis 2021; 72(5): e65–e75.

30. Bertran M, Amin-Chowdhury Z, Sheppard CL, et al. Increased Incidence of Invasive Pneumococcal Disease among Children after COVID-19 Pandemic, England. Emerg Infect Dis 2022; 28(8): 1669–72.

31. Henderson K, Gouglas D, Craw L. Gavi’s policy steers country ownership and self-financing of immunization. Vaccine 2016; 34(37): 4354–9.

32. Silaba M, Ooko M, Bottomley C, et al. Effect of 10-valent pneumococcal conjugate vaccine on the incidence of radiologically-confirmed pneumonia and clinically-defined pneumonia in Kenyan children: an interrupted time-series analysis. Lancet Glob Health 2019; 7(3): e337–e46.

33. Scott JA, Bauni E, Moisi JC, et al. Profile: The Kilifi Health and Demographic Surveillance System (KHDSS). Int J Epidemiol 2012; 41(3): 650–7.

34. Satzke C, Turner P, Virolainen-Julkunen A, et al. Standard method for detecting upper respiratory carriage of Streptococcus pneumoniae: updated recommendations from the World Health Organization Pneumococcal Carriage Working Group. Vaccine 2013; 32(1): 165–79.

35. Adetifa IMO, Bwanaali T, Wafula J, et al. Cohort Profile: The Kilifi Vaccine Monitoring Study. Int J Epidemiol 2017; 46(3): 792-h.

36. Adetifa IMO, Karia B, Mutuku A, et al. Coverage and timeliness of vaccination and the validity of routine estimates: Insights from a vaccine registry in Kenya. Vaccine 2018; 36(52): 7965–74.

37. Lucinde RK, Karia B, Ouma N, et al. The impact of the COVID-19 pandemic on vaccine coverage in Kilifi, Kenya: a retrospective cohort study. Vaccine 2022.

38. Ojal J, Flasche S, Hammitt LL, et al. Sustained reduction in vaccine-type invasive pneumococcal disease despite waning effects of a catch-up campaign in Kilifi, Kenya: A mathematical model based on pre-vaccination data. Vaccine 2017; 35(35 Pt B): 4561–8.

39. Gambia Pneumococcal Surveillance G, Mackenzie GA, Hill PC, et al. Impact of the introduction of pneumococcal conjugate vaccination on invasive pneumococcal disease and pneumonia in The Gambia: 10 years of population-based surveillance. Lancet Infect Dis 2021; 21(9): 1293–302.

40. Moore MR, Link-Gelles R, Schaffner W, et al. Effect of use of 13-valent pneumococcal conjugate vaccine in children on invasive pneumococcal disease in children and adults in the USA: analysis of multisite, population-based surveillance. Lancet Infect Dis 2015; 15(3): 301–9.

41. Pneumonia Etiology Research for Child Health Study G. Causes of severe pneumonia requiring hospital admission in children without HIV infection from Africa and Asia: the PERCH multi-country case-control study. Lancet 2019; 394(10200): 757–79.

42. Kobayashi M, Farrar JL, Gierke R, et al. Use of 15-Valent Pneumococcal Conjugate Vaccine Among U.S. Children: Updated Recommendations of the Advisory Committee on Immunization Practices – United States, 2022. MMWR Morb Mortal Wkly Rep 2022; 71(37): 1174–81.

43. Kobayashi M, Farrar JL, Gierke R, et al. Use of 15-Valent Pneumococcal Conjugate Vaccine and 20-Valent Pneumococcal Conjugate Vaccine Among U.S. Adults: Updated Recommendations of the Advisory Committee on Immunization Practices – United States, 2022. MMWR Morb Mortal Wkly Rep 2022; 71(4): 109–17.

44. European Centre for Disease Prevention and Control (ECDC). Pneumococcal Disease: Recommended vaccinations. 2023. https://vaccine-schedule.ecdc.europa.eu/Scheduler/ByDisease?SelectedDiseaseId=25&SelectedCountryIdByDisease=-1 (accessed 26 Oct 2023).

45. Mackenzie GA, Osei I, Salaudeen R, et al. A cluster-randomised, non-inferiority trial of the impact of a two-dose compared to three-dose schedule of pneumococcal conjugate vaccination in rural Gambia: the PVS trial. Trials 2022; 23(1): 71.

