## Supplemental information for "Sustained impact of 10-valent pneumococcal conjugate vaccine on invasive pneumococcal disease in Kenya, 2011-2022"

### Authors and affiliations

E Wangeci Kagucia[1], Brian M Nyamwaya[1], Gerald Ongayo[1], Mary Kaniu[1], Samuel Sang[1], Ruth Lucinde[1], Angela Karani[1], Donald Akech[1], Fredrick Odiwuor[1], Christine Mataza[2], Collins Tabu[3], Neema Mturi[1], Siti Ndaa[1], Caroline Mulunda[1], Timothy Etyang[1], Nadia Aliyan[4], Amek Nyaguara[1], Shirine Voller[1,5], Christian Bottomley[5], Laura Hammitt[6], Ifedayo Adetifa[1], J Anthony G Scott[1,5]

[1]KEMRI-Wellcome Trust Research Programme, Kilifi, Kenya
[2]Department of Health, Kilifi County, Kenya
[3]United Nations Childrens' Fund, Nairobi, Kenya
[4]Kilifi County Referral Hospital, Kilifi, Kenya
[5]London School of Hygiene and Tropical Medicine, London, UK
[6]Department of International Health, Johns Hopkins Bloomberg School of Public Health, Baltimore, US

**Table S1.** Coverage with three doses of PCV among children aged 12-23 months within the Kilifi Health and Demographic Surveillance System, 2018-2021

| Year | Number of children aged 12-23 months by 15th July | Number received 3 doses of PCV3 | % Vaccination coverage (95%CI) |
|---|---|---|---|
| **2018** | 191 | 146 | 76.4 (69.8-82.2) |
| **2019** | 157 | 140 | 89.2 (83.2-93.6) |
| **2020** | 253 | 232 | 91.7 (87.6-94.8) |
| **2021** | 413 | 370 | 89.6 (86.2-92.4) |

**Table S2.** Crude and adjusted incidence rate ratios (IRR) comparing pre- and post-vaccine invasive pneumococcal disease (IPD) incidence among residents of the Kilifi Health and Demographic Surveillance System. Significance values for the test of equality in coefficients in the late post-vaccine and COVID-19 periods *vs* the early post-vaccine period are also provided.

| | **Early post-vaccine period (2012-2016)** | | **Late post-vaccine period (2017-2019)** | | | **COVID-19 period (2020-2022)** | | |
|---|---|---|---|---|---|---|---|---|
| | **Crude IRR (95% CI)** | **Adjusted IRR* (95% CI)** | **Crude IRR (95% CI)** | **Adjusted IRR* (95% CI)** | **Late *vs* early post-vaccine period coefficient p-value** | **Crude IRR (95% CI)** | **Adjusted IRR* (95% CI)** | **COVID-19 *vs* early post-vaccine period coefficient p-value** |
| **IPD caused by any serotype** | | | | | | | | |
| <2 months | 0.03 (0.00-0.25) | 0.14 (0.01-1.27) | 0.08 (0.01-0.66) | 0.66 (0.06-7.56) | 0.474 | 0.21 (0.05-0.85) | 2.70 (0.35-20.97) | **0.029** |
| <5 years | 0.18 (0.12-0.29) | 0.32 (0.17-0.58) | 0.15 (0.08-0.29) | 0.33 (0.14-0.79) | 0.990 | 0.10 (0.05-0.20) | 0.26 (0.09-0.70) | 0.847 |
| 5-14 years | 0.34 (0.17-0.66) | 0.47 (0.25-0.89) | 0.26 (0.10-0.66) | 0.54 (0.20-1.41) | 0.953 | 0.09 (0.03-0.29) | 0.19 (0.06-0.63) | 0.274 |
| >=15 years | 0.56 (0.33-0.96) | 0.64 (0.37-1.10) | 0.26 (0.11-0.60) | 0.45 (0.17-1.20) | 0.713 | 0.09 (0.03-0.30) | 0.22 (0.05-0.94) | 0.245 |
| **IPD caused by serotype in PCV10** | | | | | | | | |
| <2 months | | | 0.11 (0.01-0.94) | 1.28 (0.09-19.01) | | | | |
| <5 years | 0.05 (0.02-0.12) | 0.08 (0.03-0.22) | 0.07 (0.03-0.19) | 0.14 (0.04-0.49) | 0.624 | 0.01 (0.00-0.09) | 0.03 (0.00-0.25) | 0.548 |
| 5-14 years | 0.19 (0.08-0.44) | 0.24 (0.11-0.56) | 0.17 (0.06-0.54) | 0.32 (0.09-1.08) | 0.683 | | | |
| >=15 years | 0.16 (0.06-0.45) | 0.19 (0.07-0.51) | 0.11 (0.03-0.50) | 0.20 (0.04-1.01) | 0.925 | | | |
| **IPD caused by serotype not in PCV10** | | | | | | | | |
| <2 months | 0.12 (0.02-0.91) | 0.30 (0.02-4.00) | | | | 0.80 (0.23-2.83) | 4.11 (0.20-84.17) | **0.043** |
| <5 years | 0.58 (0.36-0.93) | 1.27 (0.63-2.56) | 0.39 (0.19-0.81) | 1.21 (0.42-3.44) | 0.989 | 0.36 (0.18-0.73) | 1.40 (0.44-4.42) | 0.967 |
| 5-14 years | 1.10 (0.54-2.22) | 1.55 (0.72-3.35) | 0.67 (0.23-1.96) | 1.58 (0.44-5.66) | 0.999 | 0.53 (0.18-1.54) | 1.30 (0.37-4.53) | 0.947 |
| >=15 years | 1.36 (0.64-2.91) | 1.52 (0.69-3.35) | 0.56 (0.19-1.65) | 0.92 (0.24-3.51) | 0.622 | 0.27 (0.07-0.99) | 0.62 (0.10-3.71) | 0.458 |
| **Bacteraemic pneumococcal pneumonia caused by any serotype** | | | | | | | | |
| <5 years | 0.20 (0.11-0.35) | 0.15 (0.07-0.33) | 0.17 (0.08-0.38) | 0.11 (0.03-0.35) | 0.791 | 0.14 (0.06-0.32) | 0.09 (0.02-0.31) | 0.485 |
| 5-14 years | 0.38 (0.18-0.84) | 0.48 (0.21-1.06) | 0.33 (0.11-0.95) | 0.53 (0.16-1.74) | 0.982 | 0.05 (0.01-0.39) | 0.09 (0.01-0.75) | 0.236 |
| **Pneumococcal meningitis caused by any serotype** | | | | | | | | |
| <5 years | 0.08 (0.02-0.29) | 0.31 (0.08-1.20) | 0.05 (0.01-0.43) | 0.37 (0.04-3.38) | 0.986 | 0.14 (0.04-0.50) | 1.35 (0.26-6.96) | 0.167 |
| 5-14 years | 0.21 (0.06-0.70) | 0.33 (0.10-1.04) | 0.20 (0.04-1.03) | 0.54 (0.10-2.89) | 0.832 | 0.15 (0.03-0.76) | 0.42 (0.08-2.23) | 0.955 |

*IRR adjusted for year for the <2 months and <5 years age groups, and blood culture collection for the 5-14 years and ≥15 years age groups
